# Forecasting the spread of COVID19 in Hungary

**DOI:** 10.1101/2020.11.19.20234815

**Authors:** Owais Mujtaba Khanday, Samad Dadvandipour, Mohd. Aaqib Lone

**Affiliations:** Institute of Information Science, University of Miskolc, Miskolc, Hungary

**Keywords:** COVID19, Auto Regression, Exponential Smoothing, SIR Model, Holt Method

## Abstract

Time series analysis of the COVID19/ SARS-CoV-2 spread in Hungary is presented. Different methods effective for short-term forecasting are applied to the dataset, and predictions are made for the next 20 days. Autoregression and other exponential smoothing methods are applied to the dataset. SIR model is used and predicted 64% of the population could be infected by the virus considering the whole population is susceptible to be infectious Autoregression, and exponential smoothing methods indicated there would be more than a 60% increase in the cases in the coming 20 days. The doubling of the number of total cases is found to around 16 days using an effective reproduction number.

## Introduction

The novel coronavirus (COVID19/SARS-CoV-2) illness started in December 2019 in Hubei, Province, China. It is officially declared a pandemic by WHO on 11 March 2020. The virus has infected almost the whole globe, including Hungary and every European country. In the entire world, in just ten months until 28 October 2020, 44,236,745 are infected by this deadly virus, out of which 1171308 people have died. A global alarm may be caused by the confusion surrounding an unknown, novel coronavirus, leading a Harvard professor to suggest that 40-70 percent of the worldwide population may be contaminated in the coming year [1]. The virus is among the six different coronaviruses that affect the human respiratory system. The virus is transmitted by inhalation or exposure of the contaminated droplets or fomites, usually during the incubation period of between 2 and 14 days [2]. COVID-19 is dramatically affecting our daily life, and the economy has led to a significant scientific interest in this novel virus. Regardless of what one’s views are, we agree that, especially in high-risk situations, forecasts and their related uncertainty can and should be an essential part of the decision-making process. In addition to major public health issues, the threats levied on global supply chains and the economy as a whole is also essential. [^2, 3^]. A lot of crucial questions about this pandemic are still unanswered [4]. Apart from medicine, COVID-19 also draws attention to the field of epidemiology and statistics. The main focus in statistics lies in time series analysis and forecasting [5-7]. Short term forecasting methods have proven to be useful and are used by various researchers [8,9]. With the help of a precise prediction of the further course of development, essential countermeasures can be taken in the area of risk management and communication. The first two cases of COVID19 in Hungary were reported on 4 March. As of 28 October 2020, there have been 63,643 cumulative *confirmed cases* and 1535 *total deaths* in Hungary. To control the pandemic, the government declared a state of emergency within a week. By the end of March 2020, the lockdown was imposed, which was lifted on 18 May 2020 in Budapest and surrounding counties. After the first lockdown, the number of total confirmed cases daily increased slightly till the end of September, as seen in Fig.1 but started to grow at a tremendous rate after that, which indicates the second wave of COVID-19 in Hungary. The data is retrieved from the Centre for Systems Science and Engineering (CSSE) at Johns Hopkins University (https://github.com/CSSEGISandData/COVID-19)-accessed on 27 October 2020 [10]. The simulation and analysis are done using python. Deciding to discard, and act conservatively on any formal, statistical forecasts still implies an underlying forecasting mechanism, even though this process is not formalized. We used univariate time series models in this exercise, which presume that the data is correct, and past trends will continue to apply (including precautionary measures). Changes in the observed trends and the need for additional actions and steps in the case of negatively biased forecasts should be correlated with large, reliable forecast errors (potentially stretching beyond the prediction intervals). We plotted the graphs for *total cases, total deaths, total recovered, active cases, daily cases, daily deaths, daily recovered, maturity rat*e, and *recovery rate*. Three categories of data are given in the dataset; these are *total cases, total deaths*, and *total recovered*. The rest were calculated from the datasets in the following way. The data is calculated for each day.

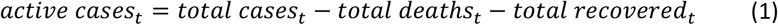

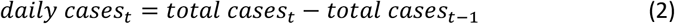

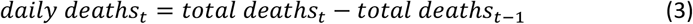

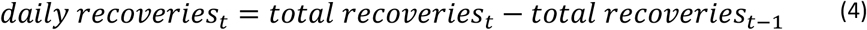

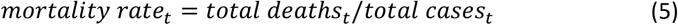

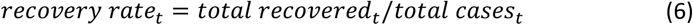

We focus on the cumulative *daily cases, total deaths, and totally recovered* across the country Hungary. The time-series graph of all these categories is in Fig. 1. By the end of June, Hungary was among the top countries in Europe to control the spread of pandemic because of the early lockdowns imposed by the government. We see the total number of cases starts growing exponentially by the end of August and is still growing exponentially. There could be many reasons for that exponential growth, two of which we think are 1) After the lockdown ended, people started to travel to different countries for the holidays. The virus could have contaminated many people as the airports, and crowded places are the epicentres of this pandemic. 2) the academic session started in September, and many international students from foreign countries travelled to Hungary for studies. Again, these students had to pass through the airports, which are highly contaminated. For theoretical solutions and optimal outcomes to different complex problems, statistical analysis of infectious diseases has played a crucial role [11]. SIR model is a well-known epidemiological model given by Kermack and McKendrick in 1927.[12] The model calculates the number of individuals infected with an epidemic in a closed population. The whole population is divided into three compartments; the susceptible S - who are vulnerable to infection, the infectious I - who have already contracted the infection and can transmit it to the susceptible population, and the recovered R - who have either died or have recovered from the disease [13]. A systematic diagram for the categories is shown in Fig 2, where *β* is the transmission rate, and *γ* is the recovery rate.

**Fig. 1.**
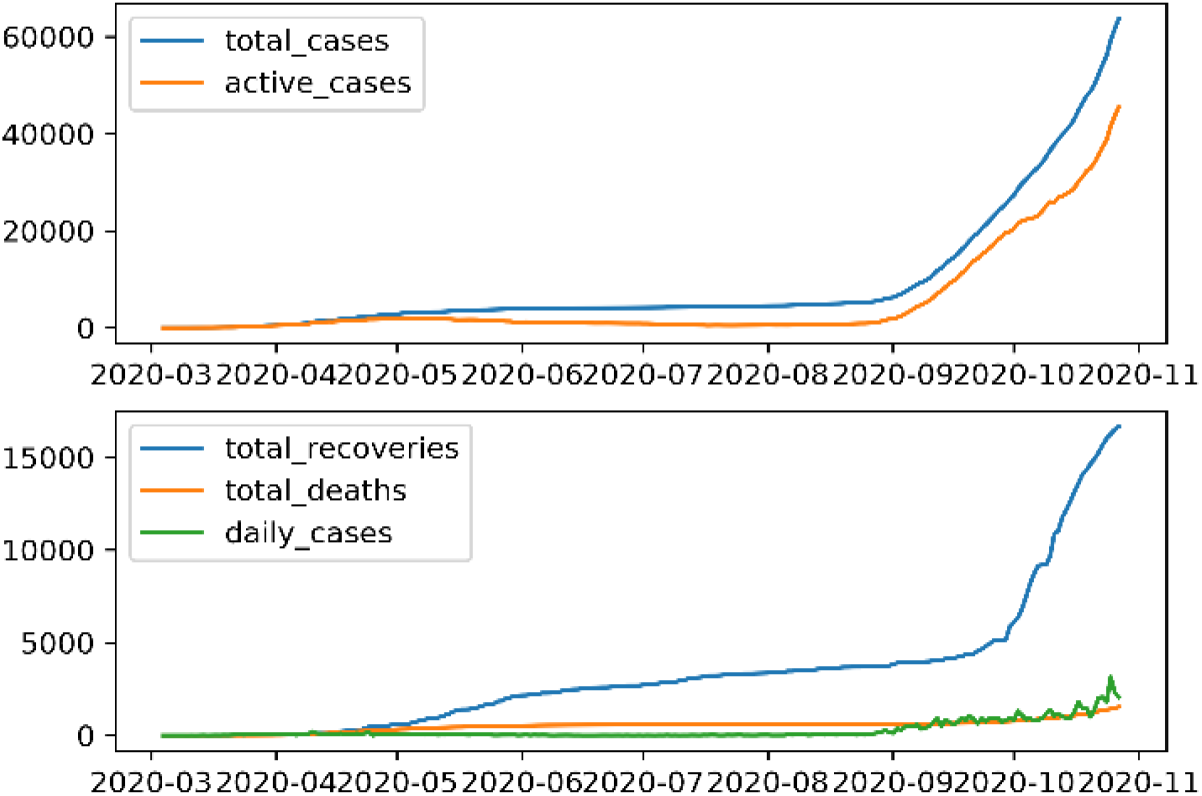
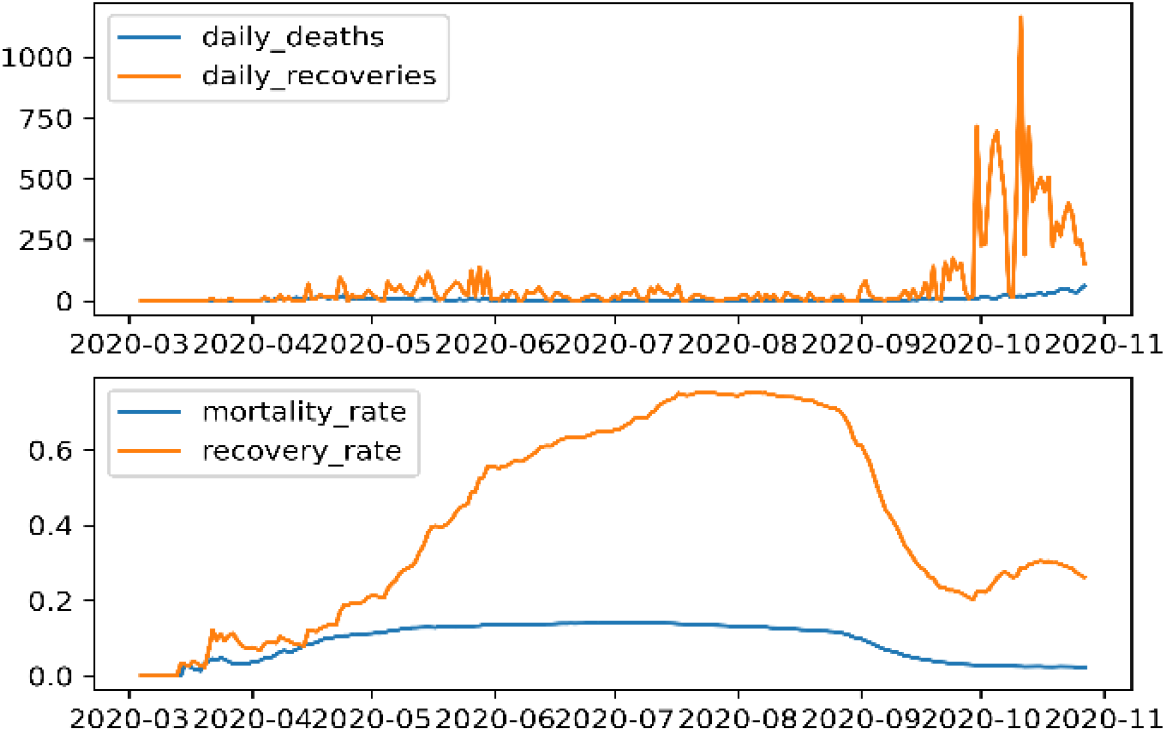
Time-series graphs of various categories starting from 03 March 2020 till 27 October 2020.

**Fig. 2.**
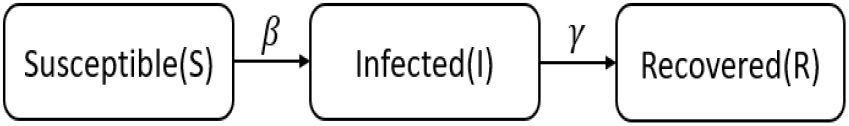
Systematic diagram for SIR Model.

The following equations describe the model:

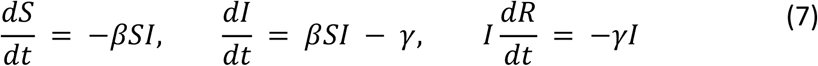

We plotted the SIR model on the dataset, as shown in Fig 2. The infection rate is taken as 3.2, and the recovery rate is calculated from the dataset, which is 0.37 (mean). The SIR model predicted more than 60% of the population will get infected if the whole population is classified as suspected. The scale is 100 for the population. We used Epiestim to calculate *R*(*t*) real-time reproduction number using Epiestim and is plotted in Fig 4. Sliding time of 7 with the preloaded dataset of Rotavirus and Gama SI distribution is used. The value of *R*(*t*) = 1.12 with a 95% confidence interval on 27 October. Using *R*(*t*), the effective doubling time of pandemic is calculated as around 16 days

The univariate analysis of the *mortality rate* and recovery rate is given in Table I. In Hungary, the *mortality rate* on average is 0.113, and the *recovery rate* is 0.372. The recovery rate is pretty good as we consider the age factor of the country’s population. The high mortality rate is high, but considering the median age of 42.7 years, this is not bad [14].

**Table I.**
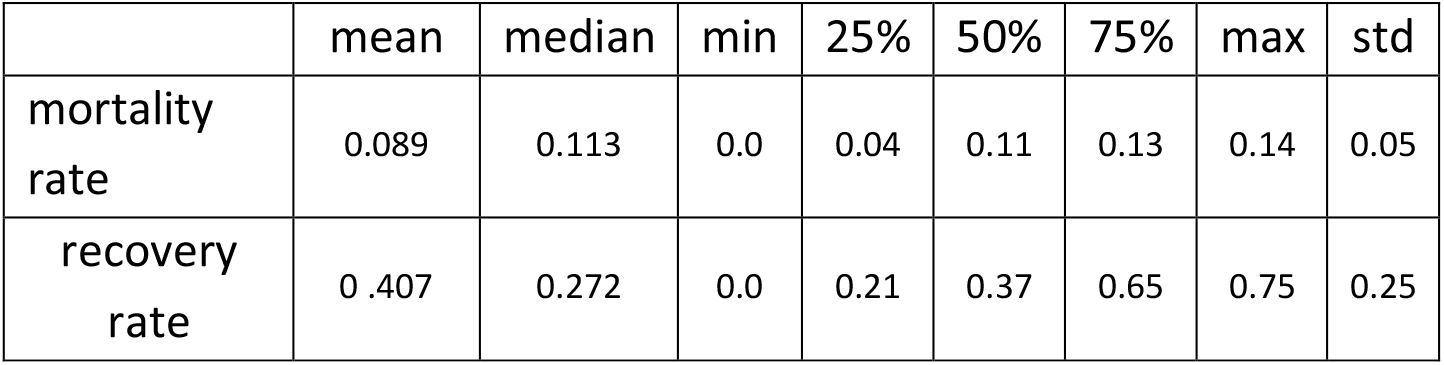
Univariate analysis of *mortality rate* and *recovery rate*.

### Analysis and Forecast using Holt exponential smoothing

Our variables of interest are *total confirmed cases, total deaths* and *total recoveries* daily. The snapshot of the data is in Fig. 1. All the variables show a slight exponential increase in April, which confirms the first wave in Hungary and no trend is observed until the end of August. After August there is a significant exponential trend which implies the second wave of COVID19 Hungary. To forecast, we adopted a family of autoregression method, exponential smoothing methods and moving average approach [15,16]. The exponential models try to capture the tread and seasonality in the data to predict the Forecast. As seen in the data, we observe there is no seasonality component involved, so our primary interest is with the trend. These methods have shown good forecast accuracy as verified by various researchers but for only short-term forecasting [17-18]. Holts exponential smoothing method with additive damp is applied to the data for the Forecast. The smoothing level and smoothing trend values applied are 0.8 and 0.2, respectively. The analysis and forecasting are done in intervals of length 20 days, starting from 17 July to 27 October. For each interval, the model is trained on the data points starting from the first reported case, i.e. 04 March to the start of the interval. The prediction is made for the next 20 days, as shown in Fig 4. We plotted the intervals to check how good the model performs on the training data.

Fig 5 shows actual data points and the predicted for all the three intervals using exponential smoothing. All the actual values lie in the range of Holt exponential trend and Additive damped trend in all the intervals except for the third interval, which starts by the end of August. The second plot of Fig 3 shows the predictions for the next 20 days. In the rest of the interval, the actual is between the additive damped and exponential trend, which implies the trend is decaying, and in the coming days, the number of cases is going to lie between the additive damped and exponential curve. The expected number of *total cases* by the end of the next 20 days is likely to increase to 110000 to 130000 as predicted by the exponential smoothing methods. The additive damped trend method predicts there will be around 100000 total cases by the end of 17 November.

**Fig. 3.**
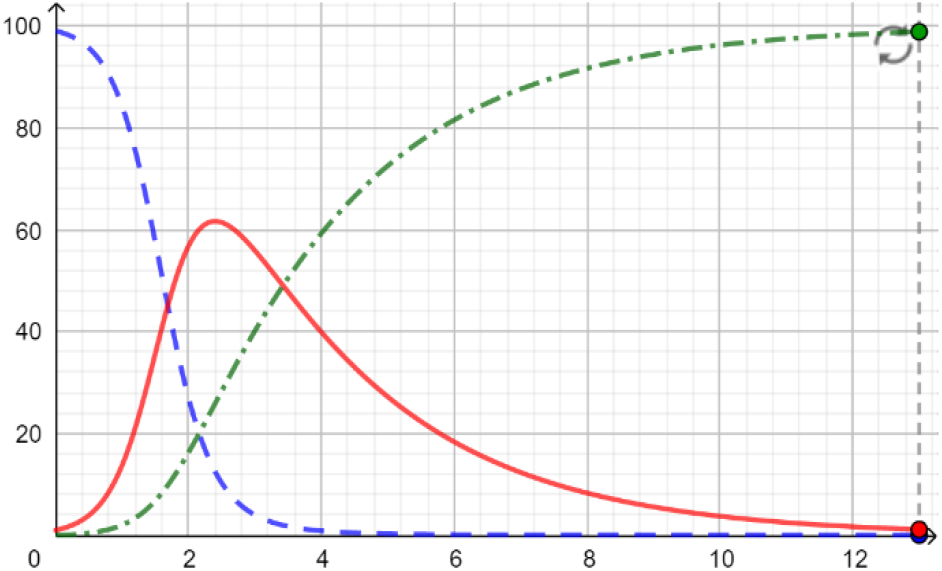
SIR Model for Hungary.

**Fig. 4.**
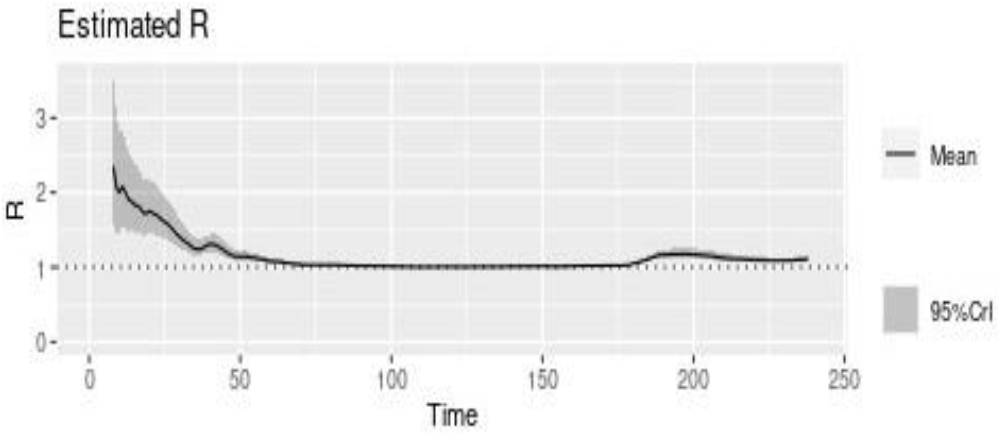
R(t) with time using Epiestim.

**Fig. 5.**
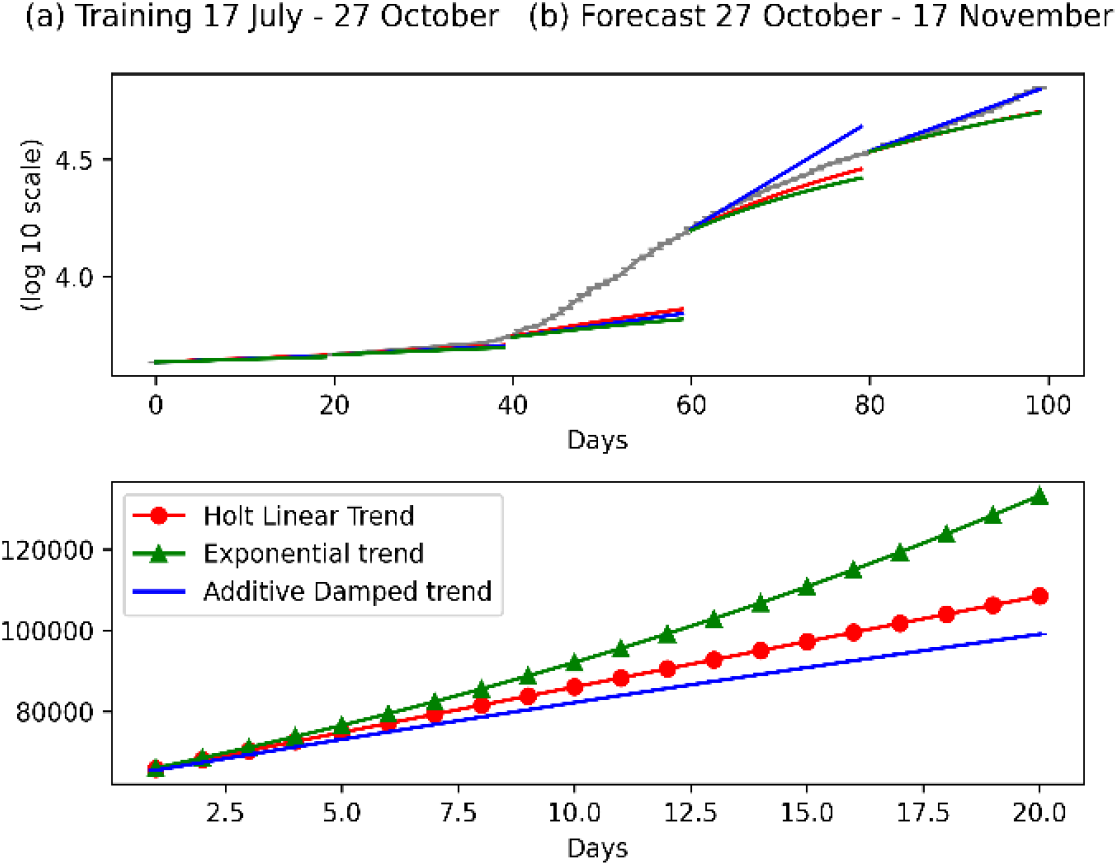
Interval plotting of *total confirmed* cases using exponential smoothing.

Fig.6 contains the plots for the *total deaths* for the same intervals as in *total cases*. The exponential smoothing models don’t perform well in any of the training data. In fact, the actual data points are more than the predicted ones. The growth rate of the deaths is more than expected by these models. The second plot gives the prediction of *total deaths* for the coming 20 days, and according to exponential smoothing models, it will be between 2200 to 2600 approximately. But in the actual, it will be more than that, as seen in the training set.

Fig 7 contains the plots for the *total recoveries* for the same intervals as in the other two categories. The exponential smoothing models don’t perform well in the 4^th^ interval of the training data. The last interval follows the additive damped trend. So in the coming next 20 days and according to the exponential smoothing model with the damped additive trend, the *total recoveries* will be between 2200 to 2400 approximately.

### Analysis and forecasting using autoregression

Autoregression models are the models that approximate non-linear time series by means of different linear autoregressive models fitted to a subset of the data. The Forecast is done using a linear combination of *past values of the variable. An autoregressive model of order p is mathematically written as*

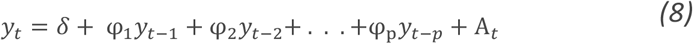

Where *y*_*t*_,*y*_t−1_,*y*_t−2_,… are the past time series values and A_t_ is the white noise and termed as randomness in the data.

The autoregression method, with a lag of 5, is applied to the *total cases* and *total deaths* dataset. A lag of 1 is applied to the total recovered dataset. The dataset was divided into training and testing sets, and the predictions for the test set are computed. The plots for these variables are shown in Fig 6.

**Fig. 6.**
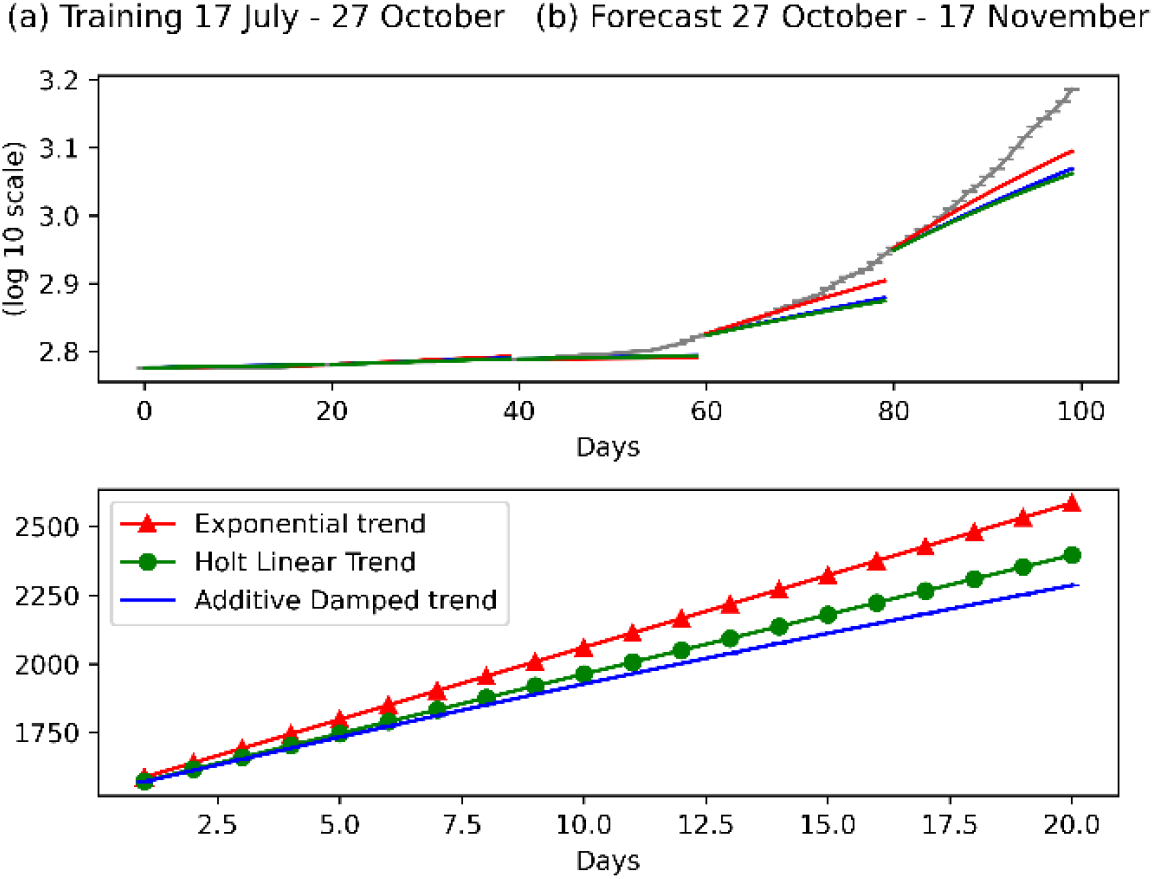
Interval plotting of *total deaths* using exponential smoothing.

The model is trained on the training data set, i.e., from 4 March to 28 September for all the variable categories. The predictions for the next 20 days are calculated and plotted in Fig 8. The predictions are compared with the actual vales, and root means the square error is calculated, as shown in Fig 8. The *total death* cases give the minimum error among the three plots. Autoregression doesn’t perform well on the *total recovered cases* test dataset.

**Fig. 7.**
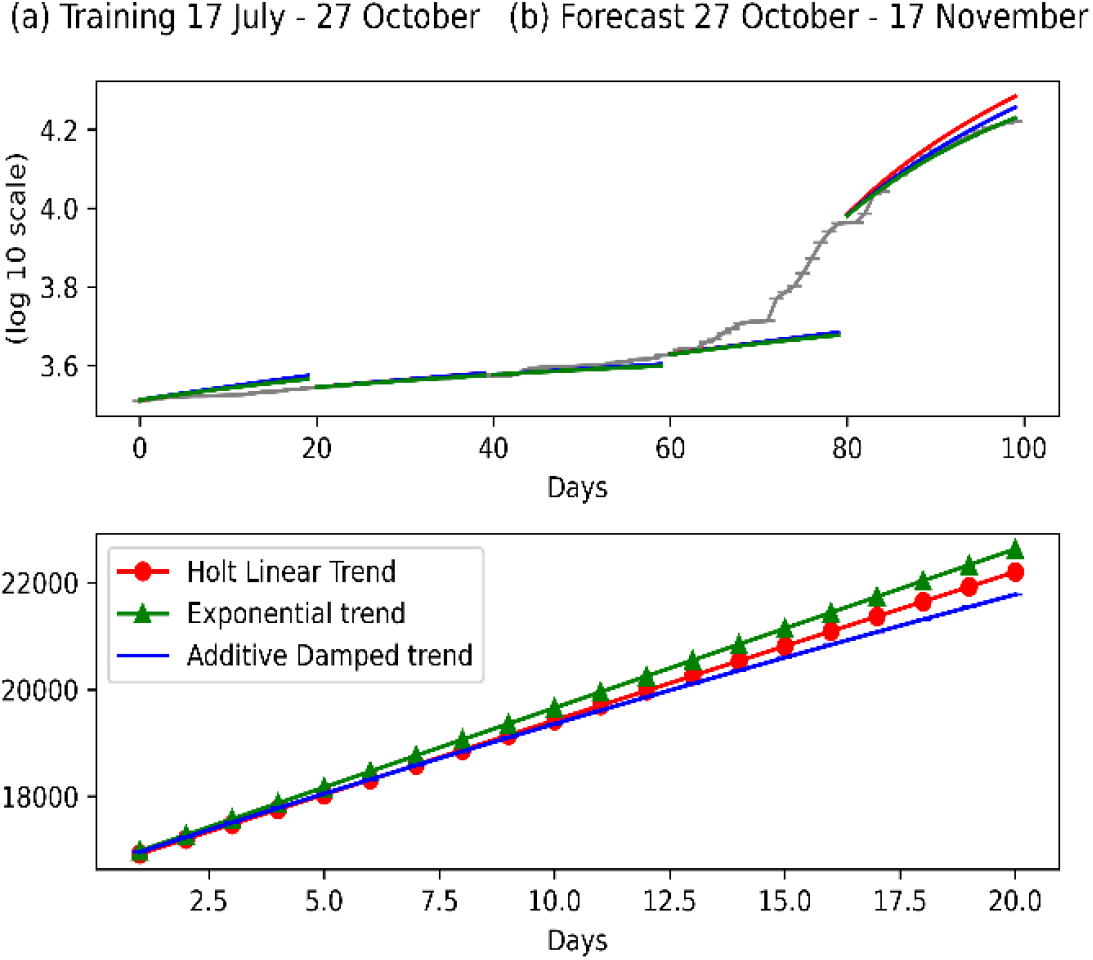
Interval plots of *total recoveries* using exponential smoothing.

**Fig. 8.**
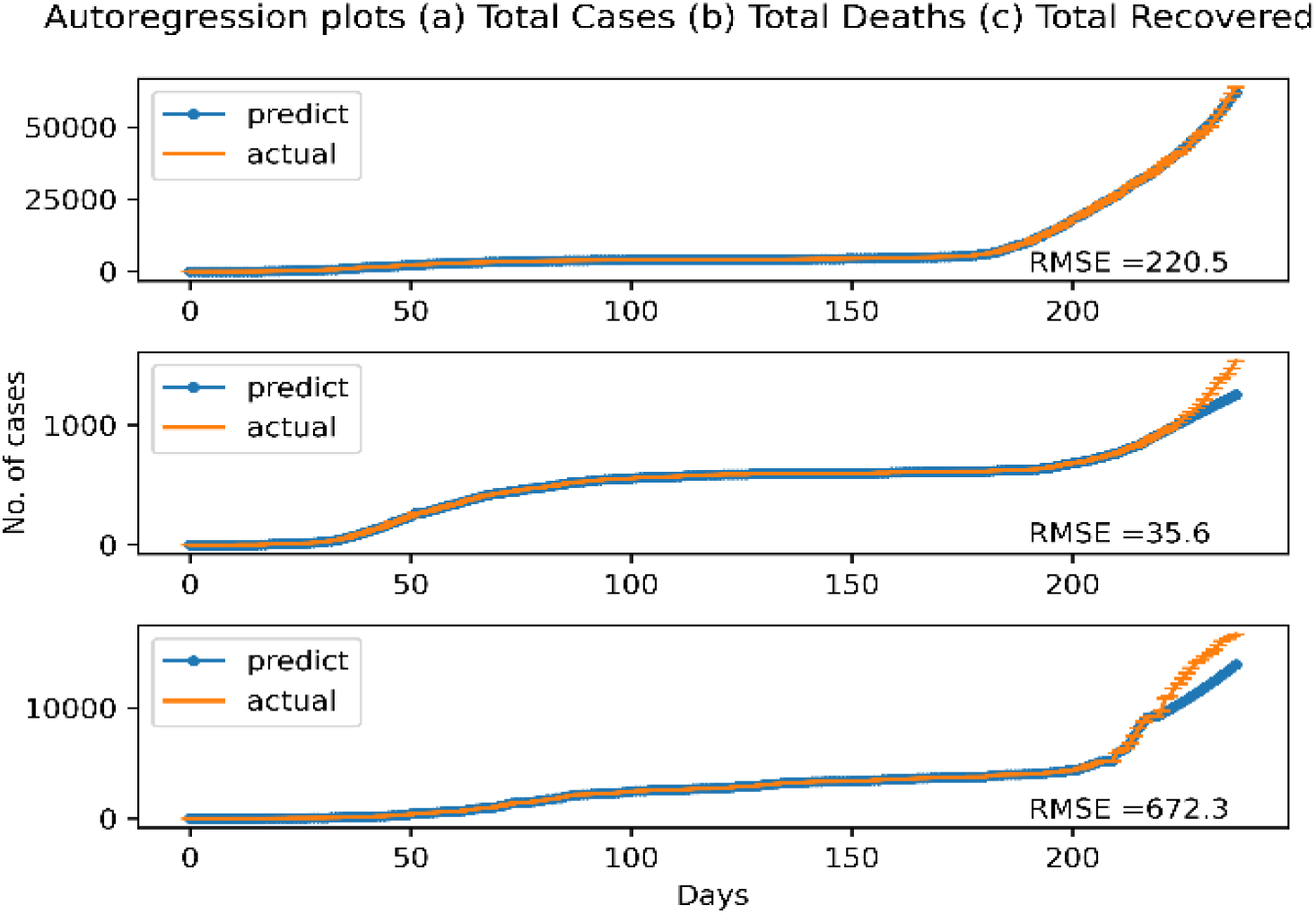
Auto Regression Plots for all three variables.

The forecasts for the next 20 day starting from the 28 October 2020 to 18 November 2020s by the autoregression is shown in Fig 9. The left plot is on a log 10 scale. We see a huge increase in the number of cases in the upcoming days. The *total recovered* curve is more increasing than the *total death* curve, which implies the recovery rate is going to increase in the upcoming days. The number of cases is increasing from 65000 to 125000 in 20 days as predicted by the autoregressive model. The numbers predicted by this model are shown in Fig 10.

**Fig. 9.**
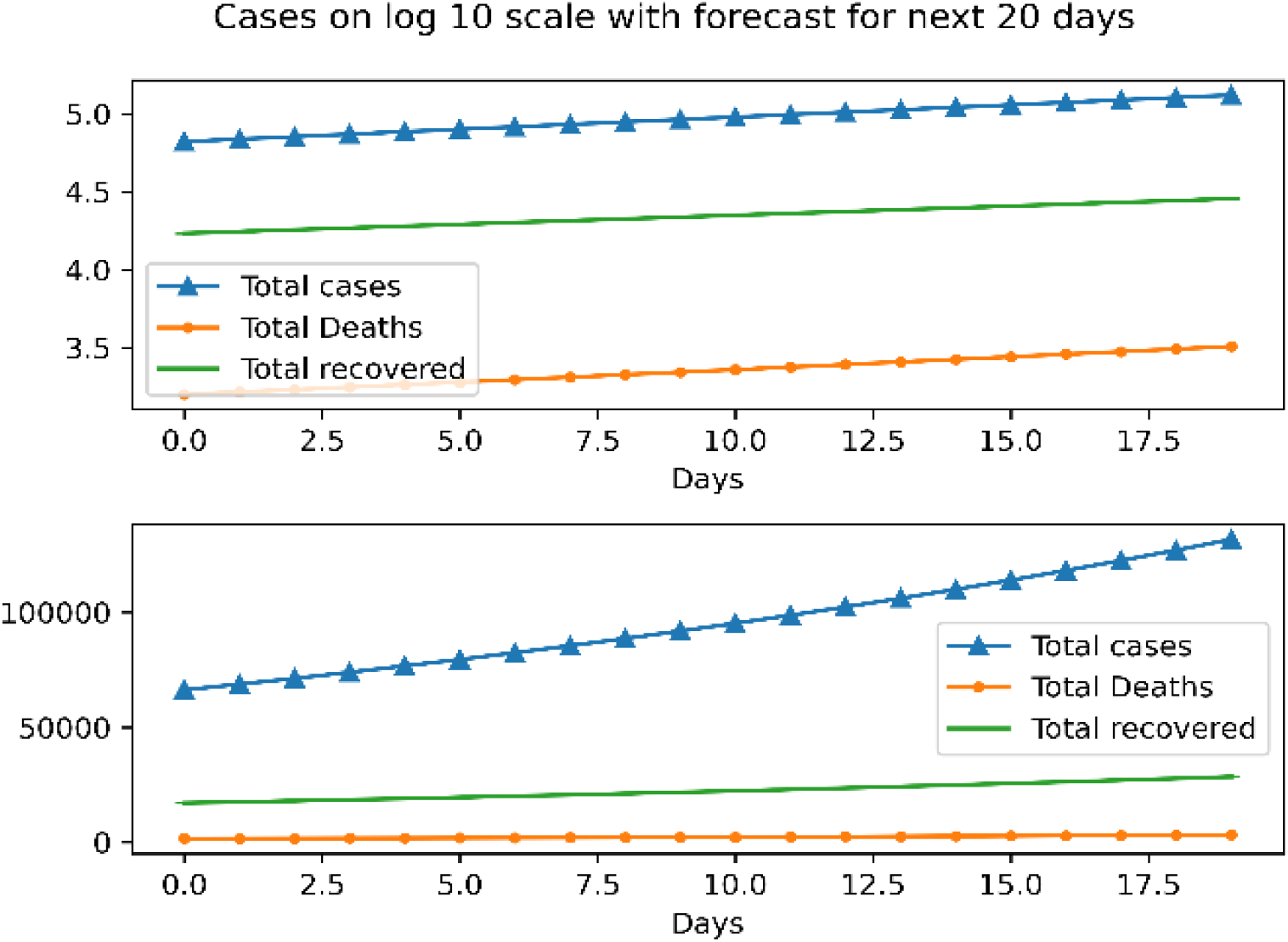
Auto Regression plot with forecasting.

**Fig. 10.**
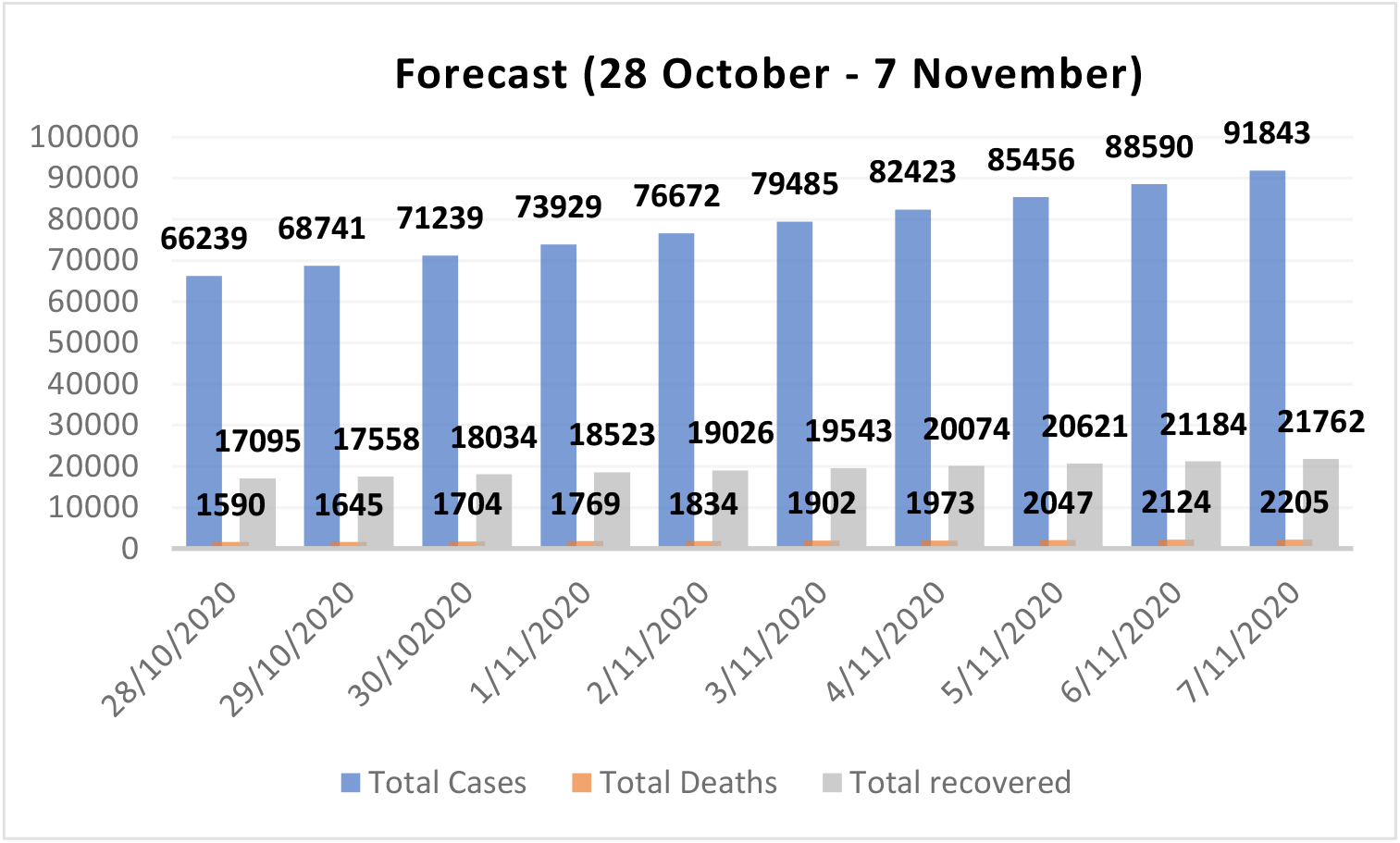
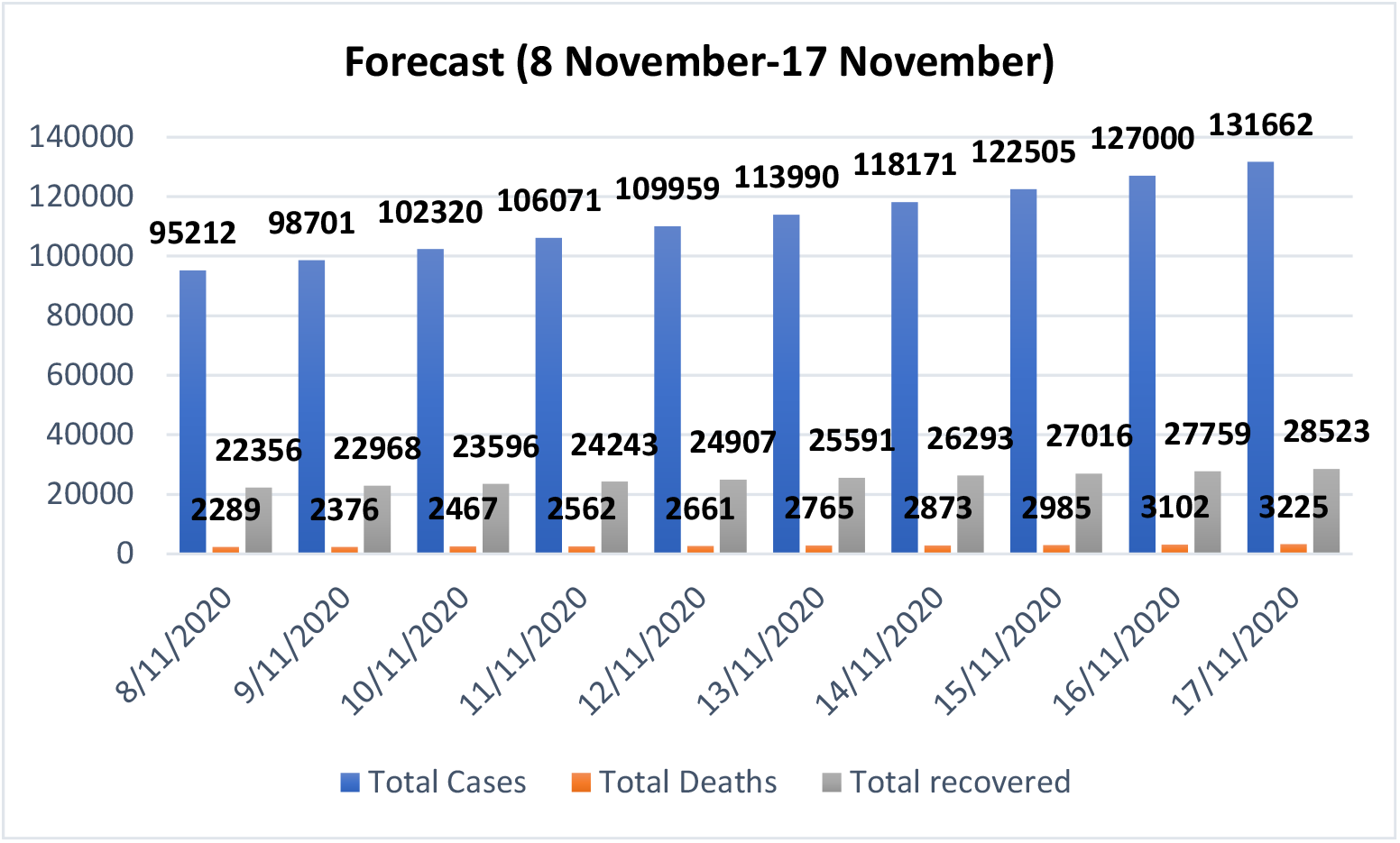
Predicted numbers for all categories using autoregression.

## Conclusion

The paper tries to predict the future spared of the COVID19 in Hungary by using the famous two models for short time series forecasting. The results of the models indicated that there would be a future exponential growth, and the number of *total cases* is going to increase from 65000 to around 130000 in the coming twenty days. Also calculated effective reproduction number revealed the cases are going to get doubled in around 16 days.

## Data Availability

COVID-19 Data Repository by the Center for Systems Science and Engineering (CSSE) at Johns Hopkins University

https://github.com/CSSEGISandData/COVID-19

## Acknowledgments

The task was performed under the framework of the EFOP-3.6.1-16-2016-00011 project, “Rejuvenating and renewing University - Innovative City of Knowledge The Institutional Development of the University of Miskolc for Intelligent Specialization.”

## Notes

### Competing Interest Statement

The authors have declared no competing interest.

### Funding Statement

EFOP 3.6.1 University of Miskolc

### Author Declarations

University of Miskolc

